# Predicting Body Composition from Chest Radiographs by Deep Learning: 10-year Mortality and Geriatric Outcomes

**DOI:** 10.64898/2026.01.13.26343990

**Authors:** Sunghwan Ji, Kiduk Kim, Kyungjin Cho, Il-Young Jang, Ji Yeon Baek, Namkug Kim, Hong-Kyu Kim, Miso Jang

**Author notes:** Corresponding author: Hong-Kyu Kim Health Screening and Promotion Center, Asan Medical Center, 88 Olympic-ro 43-gil, Songpa-gu, Seoul 05505, Republic of Korea *E-mail:* Namkug Kim, MD Department of Convergence Medicine, University of Ulsan College of Medicine, Asan Medical Center, 88 Olympic-ro 43-gil, Songpa-gu, Seoul 05505, Republic of Korea Department of Radiology and Research Institute of Radiology, University of Ulsan College of Medicine, Asan Medical Center, 88 Olympic-ro 43-gil, Songpa-gu, Seoul 05505, Republic of Korea *E-mail:. These authors contributed equally to this study (Sunghwan Ji, Kiduk Kim).

## Abstract

**Background:** Body composition strongly influences clinical outcomes in older adults, yet body mass index (BMI) lacks discriminatory power, and standard tools such as bioelectrical impedance analysis (BIA), dual-energy X-ray absorptiometry are not routinely accessible. Deep learning enables scalable, opportunistic assessment of body composition from chest radiographs (CXRs), one of the most widely available imaging modalities.

**Methods and Findings:** Using the Inception-V3 architecture, we developed a deep-learning model using 107,568 paired CXR and BIA records (2016–2018). The model was temporally validated on a separate dataset of 77,655 records (2014–2015). Our model predicted skeletal muscle mass (SMM) and fat mass (FM) with high accuracy (SMM: Pearson r = 0.967, MAE 1.40 kg; FM: r = 0.924, MAE 1.61 kg). In a cohort of 5,932 older adults (aged ≥65years), a 1-SD increase in CXR-predicted skeletal muscle index (SMI) was associated with a significant reduction in 10-year all-cause mortality (Hazard Ratio [HR] 0.65 [95% CI 0.58–0.73] for men; 0.80 [0.67–0.97] for women). In an external validation of 925 geriatric clinic patients, predicted SMI also showed comparable associations with geriatric parameters, including lower odds of sarcopenia (per 1 SD increase: 0.29 [0.22–0.38] for men; 0.25 [0.18–0.34] for women) and frailty (0.62 [0.48–0.78] for men; 1.00 [0.81–1.23] for women). These associations were more robust than those of BMI. Key limitations include the retrospective, single-center design and the use of a relatively healthy screening population.

**Conclusion:** A deep learning model applied to routine CXRs enables accurate estimation of skeletal muscle and fat mass, demonstrating prognostic and functional relevance comparable to BIA measurements. This approach may serve as a practical, low-cost tool for risk stratification and long-term care planning, particularly in older adults.

## Introduction

Accurate measurement of body composition has long been recognized as important, but practical assessment has remained challenging.^1^ One of the most widely used methods is the body mass index (BMI), which has long served as a convenient indicator of obesity.^2^ High BMI is a well-known predictor of various health-related outcomes, including all-cause and cause-specific mortality,^3^ cardiovascular diseases,^4^ several types of cancer,^5^ and disease burden.^6^ However, recent studies have shown that this association may be reversed in certain subpopulations, especially in older adults, with a J-shaped curve,^7,8^—commonly referred to as the “obesity paradox.”^9^ This paradox may stem from BMI’s inability to distinguish between body-composition components such as muscle and fat mass.^10^

Body composition is closely linked to physical performance^11^ and disability,^12^ yet this relationship is not captured by BMI^13^, especially in older adults. In the context of aging societies, accurate body-composition assessment has therefore become increasingly important. Conventional methods include bioelectrical impedance analysis (BIA), dual-energy X-ray absorptiometry (DXA), computed tomography (CT), and magnetic resonance imaging (MRI).^14^ Although BIA and DXA are less expensive than CT or MRI, they still involve additional costs and procedural steps, making them unsuitable for routine health check-ups such as national screening programs where cost-effectiveness is essential.

Recent advances in deep learning have enabled automated, opportunistic measurements of body composition.^15,16^ Some retrospective studies have shown that these opportunistic results are associated with clinical outcomes such as cancer survival,^17,18^ interventional procedural outcomes,^19^ and surgical outcomes.^20^ Nevertheless, these AI-enabled CT/MR-based approaches remain constrained by cost, limited availability, and, for CT, ionizing radiation, making routine use in primary care challenging.

Given their ubiquity and relatively low cost and radiation exposure compared with CT or DXA, chest radiographs (CXRs) may offer a scalable alternative for opportunistic body-composition assessment. Recent studies have shown that deep learning can extract biologically meaningful information directly from CXRs, such as osteoporosis risk,^21^ pulmonary function,^22^ and biologic age.^23^ Building on these advances, we developed a deep-learning model to estimate body composition from CXRs and evaluated its accuracy and prognostic value for 10-year all-cause mortality among adults aged ≥65 years in a temporally distinct validation dataset. We further examined whether CXR-based estimates demonstrated similar associations with physical performance, grip strength, sarcopenia, disability, and frailty as BIA-based measurements, using data from comprehensive geriatric assessments (CGAs) conducted in an external geriatric clinic dataset.

## Methods

This study was conducted in accordance with the current scientific guidelines and the principles of the Declaration of Helsinki. The study protocol was approved by the Institutional Review Board of Asan Medical Center, Seoul, Korea (approval no. 2022-0824). The protocol for the CGA Dataset was separately approved (approval no. 2023-1619). The requirement for informed consent was waived owing to the retrospective nature of the study.

### Study Design and Population

This study comprised three main steps: (1) development of a deep-learning model to predict body composition from CXRs (development dataset); (2) validation using a temporally distinct dataset in the same institute of development dataset (temporal validation dataset); and (3) external validation using CGA dataset (**Figure 1**).

**Figure 1.**
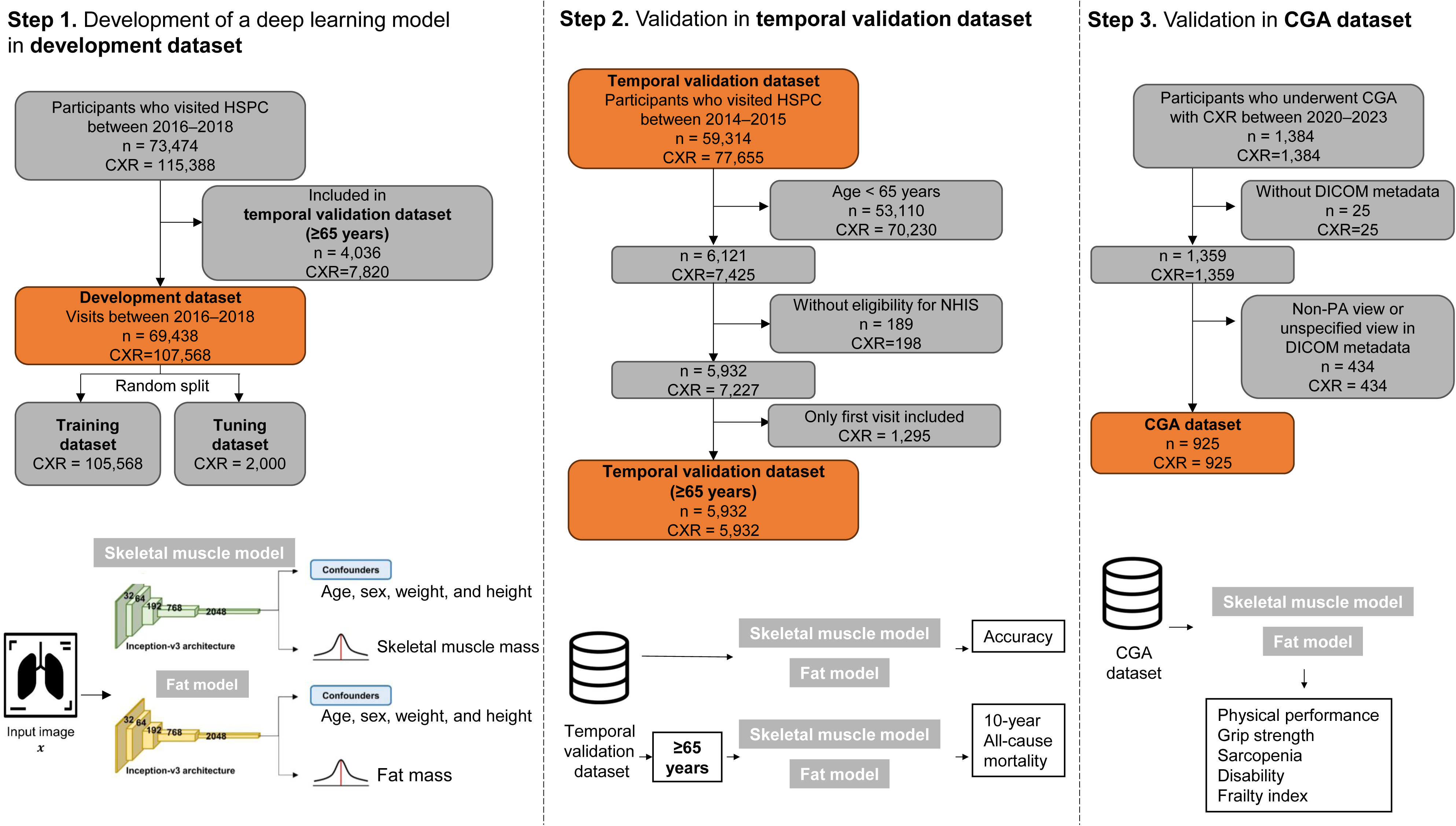
Development and Validation of Deep Learning Model for Predicting Body Composition using CXRs. **Note:** n corresponds to the number of participants, and CXR corresponds the number of chest radiographs(CXR).

We retrospectively identified participants who visited the Health Screening and Promotion Center (HSPC) between 2014 and 2018 and underwent both BIA and CXR. A total of 98,794 participants with BIA results and 192,905 corresponding CXRs obtained according to standard post-anterior (PA) imaging protocol were included as the initial study population.

For temporal validation, participants with visits between 2014 and 2015 were selected (n = 59,314; CXRs = 77,655) for accuracy analyses. After excluding those younger than 65 years (n = 53,110; CXRs = 70,230) and those without eligibility in the National Health Insurance Service (NHIS) database (n = 189; CXRs = 198), 5,932 older adults (CXRs = 5,932) were included in the temporal validation dataset (≥65 years) for mortality analyses. Participants with visits between 2016 and 2018 (n = 73,474; CXRs = 115,388) were used for model development. Individuals included in the temporal validation dataset (≥65 years) dataset were excluded from the initial AI model development dataset to avoid data leakage. Thus, BIA and CXR data from 107,568 visits were used to develop the model, which was subsequently divided into 105,568 visits for training and 2,000 visits for tuning. The flowchart of patient selection and dataset construction is shown in **Figure 1**. Data partitions were disjoint at the visit level; thus, each visit was included in only one partition. After the data were split to prevent data leakage, the identifiers, including the names, were removed for de-identification purposes.

For external validation, we identified an independent cohort of participants from the geriatric clinic, same institution who underwent both CGA and CXR between April 1, 2020, and October 15, 2023 (N = 1,384; CXRs = 1,384). After excluding those without DICOM metadata (n = 25; CXRs = 25) and those with non-PA view or unspecified view in DICOM metadata (n = 434; CXRs = 434), a total of 925 participants were included in the CGA dataset (n = 925; CXRs = 925). This CGA dataset was used to evaluate whether CXR-based skeletal muscle mass (SMM) demonstrated similar associations with geriatric outcomes—including gait speed, physical performance, grip strength, sarcopenia, disability, and frailty index—as BIA-based measures.

### Body-composition Measurement

In development and temporal validation dataset, body composition was measured through a bioelectrical impedance analyzer (Inbody 720; InBody Co. Ltd., Seoul, Korea), which is widely used as a standard tool for body composition assessment in clinical and epidemiological studies.^24^ The device requires the patient to stand on it and hold tactile electrodes that send detectable electrical signals through the body for measuring the impedance of body tissues to estimate their segmental body composition. This device employs six frequencies (1, 5, 50, 250, 500, and 1,000 kHz) to generate 30 impedance values for five body segments. Note that the body-composition measurements were conducted after overnight fasting.

This study employed the body compositions of SMM and fat mass (FM). The SMM was adjusted using the square of the patient’s height (skeletal muscle index, SMI), whereas fat percentage was defined as the FM in kilograms divided by their weight (%). The CXR-predicted body composition was adjusted using the predicted height and weight.

### Model Development

The deep-learning model for predicting body compositions from CXRs was developed using the Inception v3 Architecture.^25^ The input CXRs were resized to 512 × 512 pixels while maintaining their original image ratio through zero padding. BIA-based body compositions, such as SMM and FM, were used as the ground truths for the predictions. Additionally, clinical information, including height and weight, was also input into the model to account for confounding variables for the multitask prediction.^26^ The model functions are detailed in **Method S1**.

### 10-year All-cause Mortality

In the temporal validation dataset (≥65 years)), all-cause mortality was assessed based on patient eligibility records from the National Health Insurance Service (NHIS). Because all South Korean citizens are enrolled in the NHIS, vital status and date of death can be accurately derived from its database. Survival status was determined up to August 1, 2023.

### Geriatric Outcomes

In the CGA dataset, CGA was conducted by trained nurses in the geriatric clinic. Patients underwent CGA for various purposes, including preoperative evaluation and baseline geriatric assessment. Gait speed and the Short Physical Performance Battery (SPPB) were measured using a one-dimensional LiDAR device within the eSPPB kit (Dyphi Inc., Daejeon, Korea).^27^ Low gait speed and low SPPB were defined as <1.0 m/s and <9 points, respectively.^28^ Grip strength was measured with a handgrip dynamometer (T.K.K. 5401 Grip-D; Takei, Tokyo, Japan), and low grip strength was defined as <28 kg for men and <18 kg for women.^28^ Body composition was measured through a bioelectrical impedance analyzer (Inbody S10; InBody Co. Ltd., Seoul, Korea). Sarcopenia was assessed according to the Asian Working Group for Sarcopenia (AWGS) guidelines.^28^

Frailty was evaluated using a 50-item Frailty Index (FI), constructed based on the deficit accumulation model and established methodology.^29^ The FI quantified accumulated health deficits using CGA parameters and was calculated as the ratio of the number of deficits present to the total number of items (0–1 scale), with higher scores indicating greater frailty. The 50 items included underlying diseases, physical and cognitive impairments, psychosocial risk factors, geriatric syndromes, and disability (**Table S1**). Frailty was defined as frailty index lower than 0.25.^30^ Disability was defined as the presence of impairment in ≥1 of 7 activities of daily living (ADL) or 10 instrumental activities of daily living (IADL).^31^

### Statistical Analysis

The mean average errors (MAEs) between the reference standard and CXR-based values were calculated for continuous variables, such as SMM, FM, age, height, and weight, whereas the accuracies were calculated for categorical variables, such as sex. Pearson’s correlation coefficient (*r*) with 95% confidence interval (CI) between the BIA-based and CXR-based body compositions was also calculated.

In the temporal validation dataset (≥65 years), hazard ratios (HRs) per 1 standard deviation (SD) increase in body composition measures were estimated for all-cause mortality using Cox proportional hazards models. Both BIA-based and CXR-predicted indices SMI and fat percentage were analyzed separately for men and women. Predicted HRs across the continuous range of each body composition measure were derived using marginal estimates, and HR–body composition curves with 95% confidence intervals (CIs) were visualized. BMI (weight divided by height squared) was also analyzed for comparison.

In the CGA dataset, odds ratios (ORs) per 1 SD increase in SMI, with corresponding 95% CIs, were calculated for low gait speed, low physical performance, low grip strength, sarcopenia ADL disability, IADL disability, and frailty. Because fat percentage showed limited associations with geriatric parameters such as frailty and disability, analyses and results were primarily focused on SMI.^32,33^

All analyses were performed using R software (version 4.2.1; R Foundation for Statistical Computing, Vienna, Austria) with the *survival*, *ggeffects*, and *epiR* packages. Statistical significance was defined as a two-sided *P* value < 0.05. Continuous variables are presented as mean ± standard deviation (SD), and categorical variables as frequencies and percentages. This study was reported in accordance with the CLAIM (Checklist for Artificial Intelligence in Medical Imaging) and STROBE (Strengthening the Reporting of Observational Studies in Epidemiology) guidelines (see **Supplementary Material**).

## Results

### Baseline Characteristics

As descried in **Table 1**, the development dataset consisted of 47,788 females (mean age 51.05±9.90 years) and 59,778 males (mean age 52.37±9.38 years). The Temporal validation dataset included 33,085 females (mean age 50.86±10.11 years) and 44,570 males (mean age 52.44±9.85 years), with an additional subset of 2,431 females (mean age 69.90±4.44 years) and 3,501 males (mean age 69.90±4.69 years) aged ≥65 years. The CGA dataset comprised 428 females (mean age 77.52±6.55 years) and 497 males (mean age 76.61±7.40 years). Across all datasets, males generally showed higher values for height, weight, skeletal muscle mass, and grip strength, while body fat percentage was consistently higher in females. The CGA dataset participants were notably older and showed different body composition patterns, including lower muscle mass and higher fat mass indices compared to the younger cohorts.

**Table 1.**
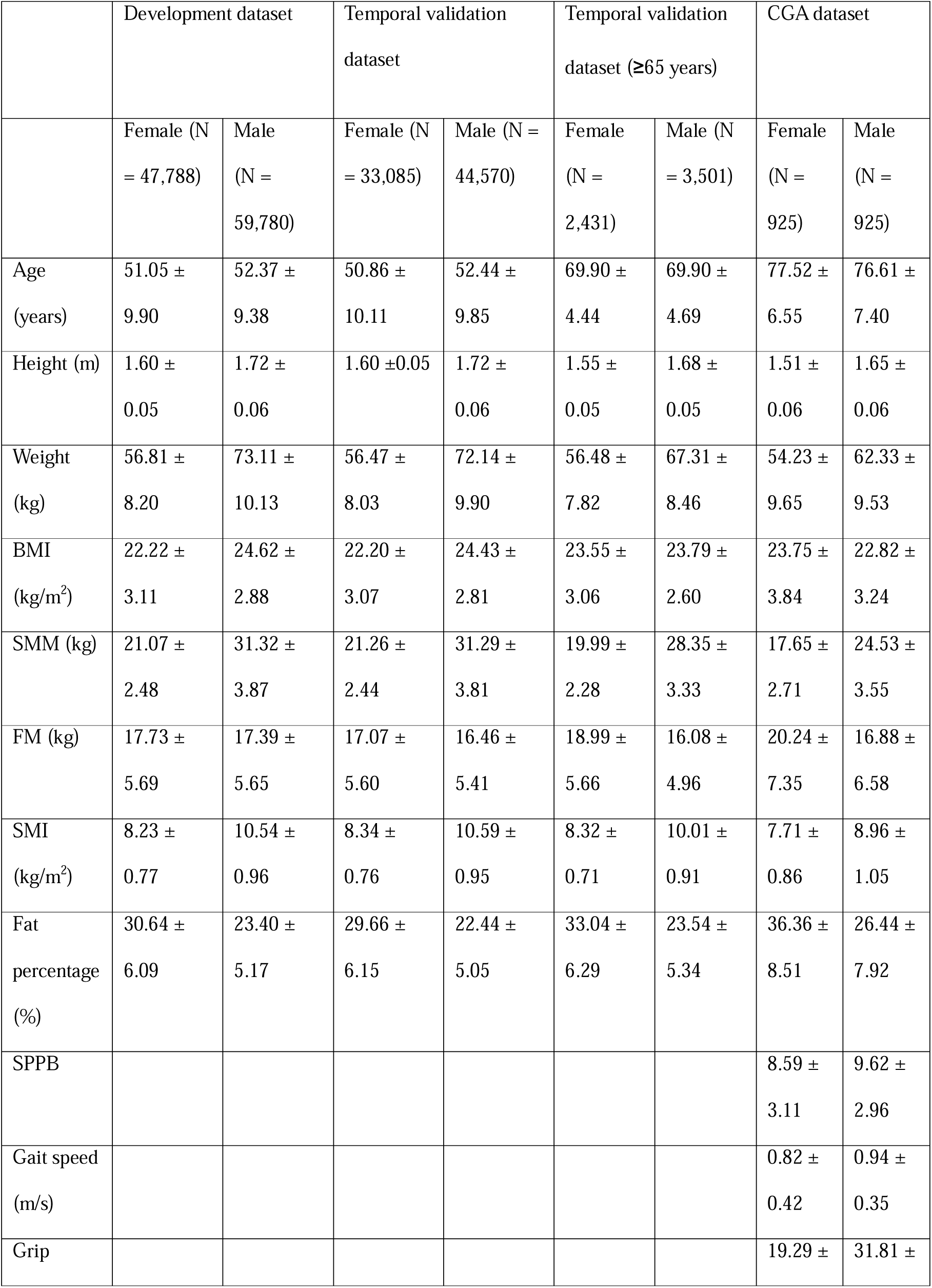
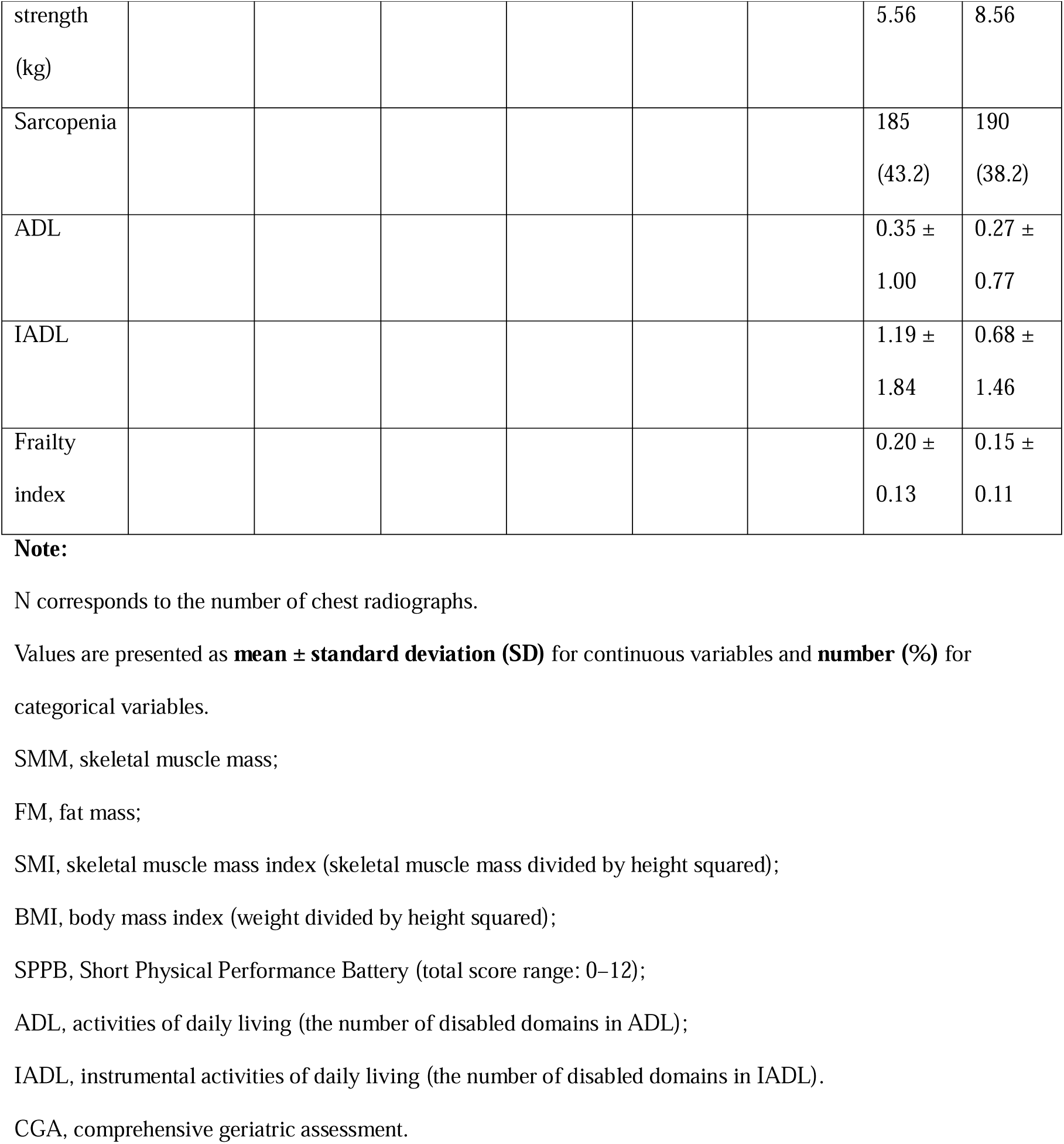
Baseline characteristics of Development, Temporal validation, and CGA dataset.

### Temporal Validation

**Table 2(A)** and **Figure. S1** present the performance results of the developed model for the temporal validation set, including the MAEs and *r* values for the SMM, FM, age, height, weight, and accuracy of sex predictions. It achieved MAE and *r* values of 1.40 ± 1.10 kg and 0.967 (0.966–0.967) for the SMM predictions, respectively, and 1.61 ± 1.37 kg and 0.924 (0.922–0.925) for the FM predictions, respectively. Additionally, the performance was similar for the temporal validation dataset (≥65 years), except for the age (see **Table S2**). The prediction accuracies across different sex subgroups were also comparable (**Tables S3 and S4**). The average grad-cam results are shown in **Figure. S2**.

**Table 2.**
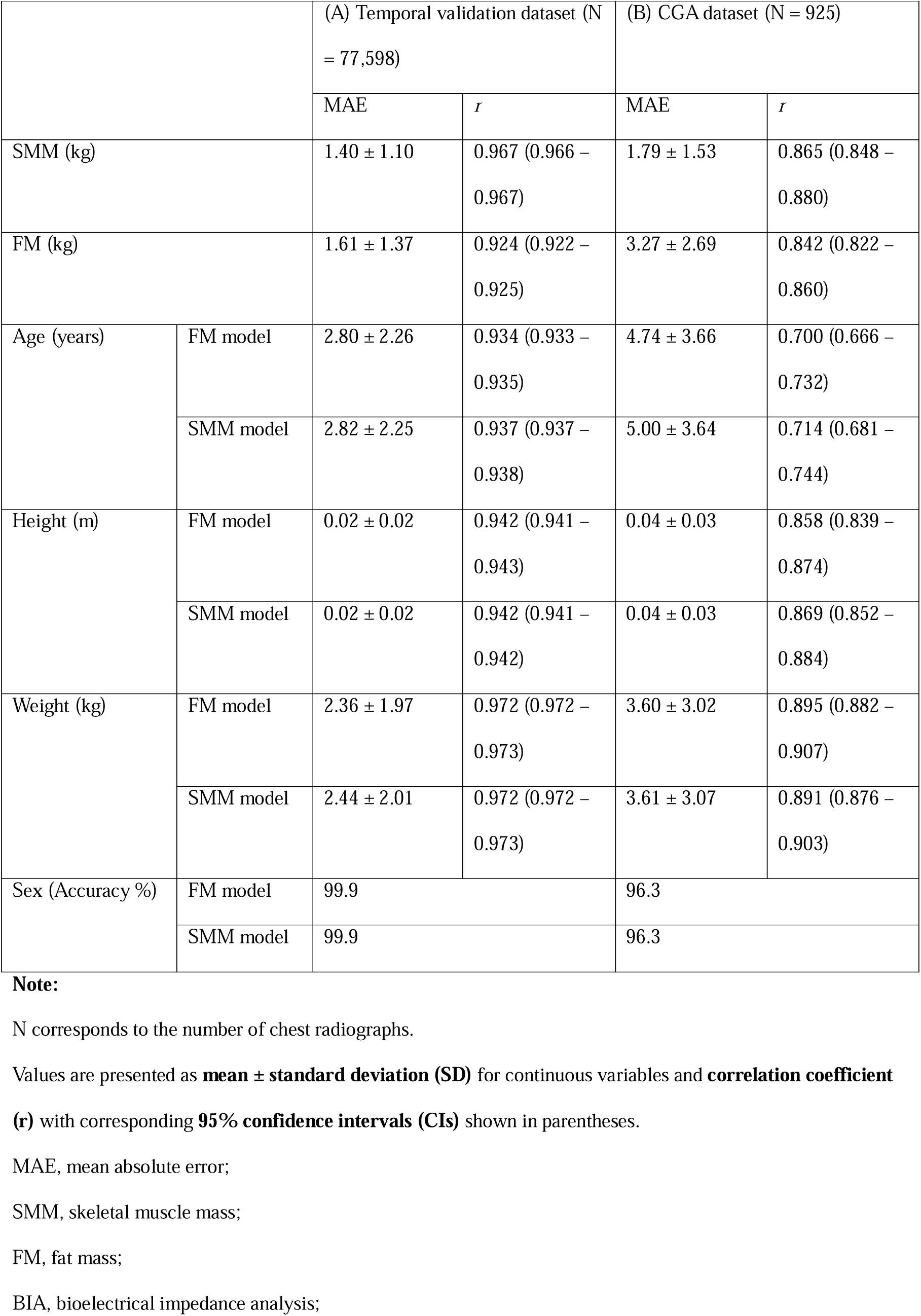
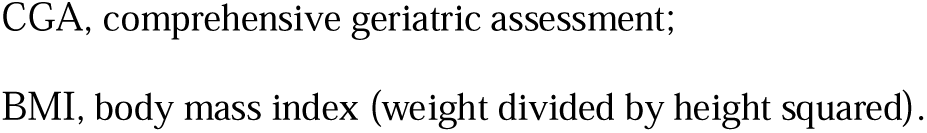
Mean Absolute Errors (MAEs) and Correlation Coefficient (*r*) Values of Skeletal Muscle Mass (SMM), Predicted Fat Mass (FM), Age, Height, Weight, and Sex for the Temporal validation and CGA dataset.

|  |  | (A) Temporal validation dataset (N = 77,598) |  | (B) CGA dataset (N = 925) |  |
| --- | --- | --- | --- | --- | --- |
|  |  | MAE | *r* | MAE | *r* |
| SMM (kg) |  | 1.40 ± 1.10 | 0.967 (0.966 – 0.967) | 1.79 ± 1.53 | 0.865 (0.848 – 0.880) |
| FM (kg) |  | 1.61 ± 1.37 | 0.924 (0.922 – 0.925) | 3.27 ± 2.69 | 0.842 (0.822 – 0.860) |
| Age (years) | FM model | 2.80 ± 2.26 | 0.934 (0.933 – 0.935) | 4.74 ± 3.66 | 0.700 (0.666 – 0.732) |
|  | SMM model | 2.82 ± 2.25 | 0.937 (0.937 – 0.938) | 5.00 ± 3.64 | 0.714 (0.681 – 0.744) |
| Height (m) | FM model | 0.02 ± 0.02 | 0.942 (0.941 – 0.943) | 0.04 ± 0.03 | 0.858 (0.839 – 0.874) |
|  | SMM model | 0.02 ± 0.02 | 0.942 (0.941 – 0.942) | 0.04 ± 0.03 | 0.869 (0.852 – 0.884) |
| Weight (kg) | FM model | 2.36 ± 1.97 | 0.972 (0.972 – 0.973) | 3.60 ± 3.02 | 0.895 (0.882 – 0.907) |
|  | SMM model | 2.44 ± 2.01 | 0.972 (0.972 – 0.973) | 3.61 ± 3.07 | 0.891 (0.876 – 0.903) |
| Sex (Accuracy %) | FM model | 99.9 |  | 96.3 |  |
|  | SMM model | 99.9 |  | 96.3 |  |

**Note:**
N corresponds to the number of chest radiographs.
Values are presented as **mean ± standard deviation (SD)** for continuous variables and **correlation coefficient (r)** with corresponding **95% confidence intervals (CIs)** shown in parentheses.
MAE, mean absolute error;
SMM, skeletal muscle mass;
FM, fat mass;
BIA, bioelectrical impedance analysis;
CGA, comprehensive geriatric assessment; BMI, body mass index (weight divided by height squared).

In the temporal validation dataset (≥65 years), the predicted body composition parameters showed mortality patterns consistent with those derived from BIA measurements and demonstrated better discriminative ability than BMI (**Figure 2**), with each 1-SD decrease in skeletal muscle mass associated with higher mortality risk (HR 0.77 [95% CI 0.64–0.92] in women; 0.58 [0.52–0.65] in men) and each 1-SD increase in fat percentage associated with increased mortality (HR 1.09 [0.91–1.30] in women; 1.25 [1.11–1.40] in men), whereas BMI showed weaker associations (HR 0.97 [0.81–1.16] in women; 0.83 [0.74–0.93] in men). As shown in **Figure S3**, lower predicted skeletal muscle index (SMI) and higher fat percentage were associated with increased all-cause mortality. The mortality patterns estimated by CXR closely paralleled those obtained from BIA and showed stronger correlations than those with BMI. Consistency between BIA- and CXR-based estimates was high across all components, with Pearson r = 1.00 and concordance correlation coefficients ranging from 0.68 to 0.98 (**Table S5**).

**Figure 2.**
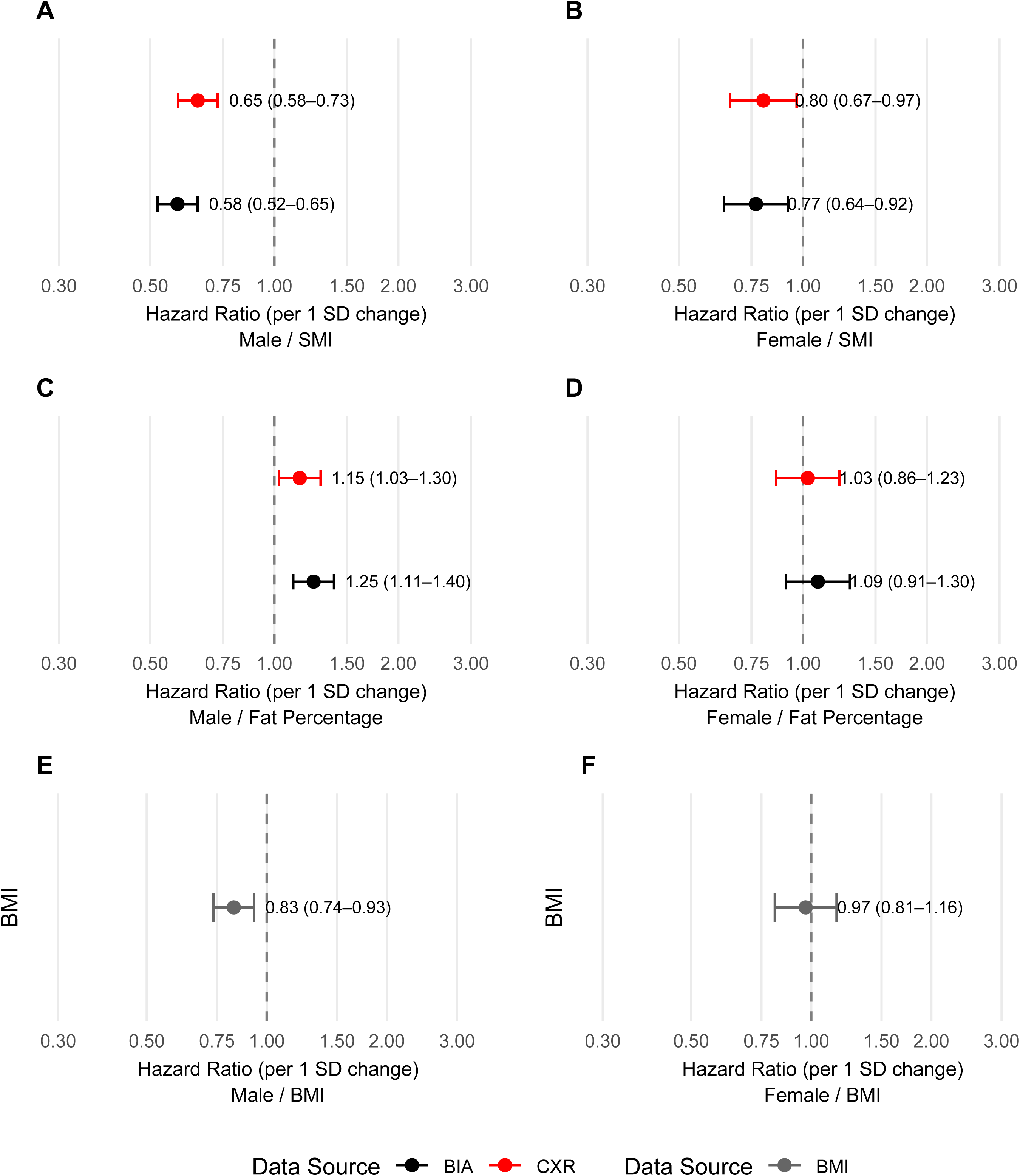
Association of body composition indices with 10-year all-cause mortality **Note:** Forest plots showing hazard ratios (HRs) per 1 standard deviation (SD) increase in skeletal muscle index (SMI), fat percentage, and body mass index (BMI) for men and women in the temporal validation dataset (≥65 years). Error bars represent 95% confidence intervals (CIs).

### CGA Validation

**Table 2(B)** presents the performance results of the developed model for the CGA dataset. Compared with the temporal validation set, the prediction performance was slightly lower across all parameters. The model achieved MAE and correlation values of 1.79 ± 1.53 kg and 0.865 (0.848–0.880) for SMM, and 3.27 ± 2.69 kg and 0.842 (0.822–0.860) for FM, respectively.

Figure 3 demonstrates that CXR-derived SMI showed patterns similar to those obtained from BIA in their associations with geriatric assessment outcomes. Both CXR- and BIA-based SMI consistently exhibited protective effects against frailty and functional impairments, with stronger associations observed in men (Figure 3A) than in women (Figure 3B), and more pronounced associations compared with those of BMI (Figure 3C **and 3D**). CXR-derived SMI was associated with lower odds of sarcopenia (per 1 SD increase: 0.29 [0.22–0.38] for men; 0.25 [0.18–0.34] for women). Although CXR-predicted SMI showed a strong association with lower odds of frailty in men (0.62 [0.48–0.78]), this association was not significant in women (1.00 [0.81–1.23]). Results for CXR-derived fat percentage are presented in **Figure S4**, showing limited associations with geriatric parameters such as disability and frailty in both BIA- and CXR-derived measures, consistent with previous findings.^32,33^

**Figure 3.**
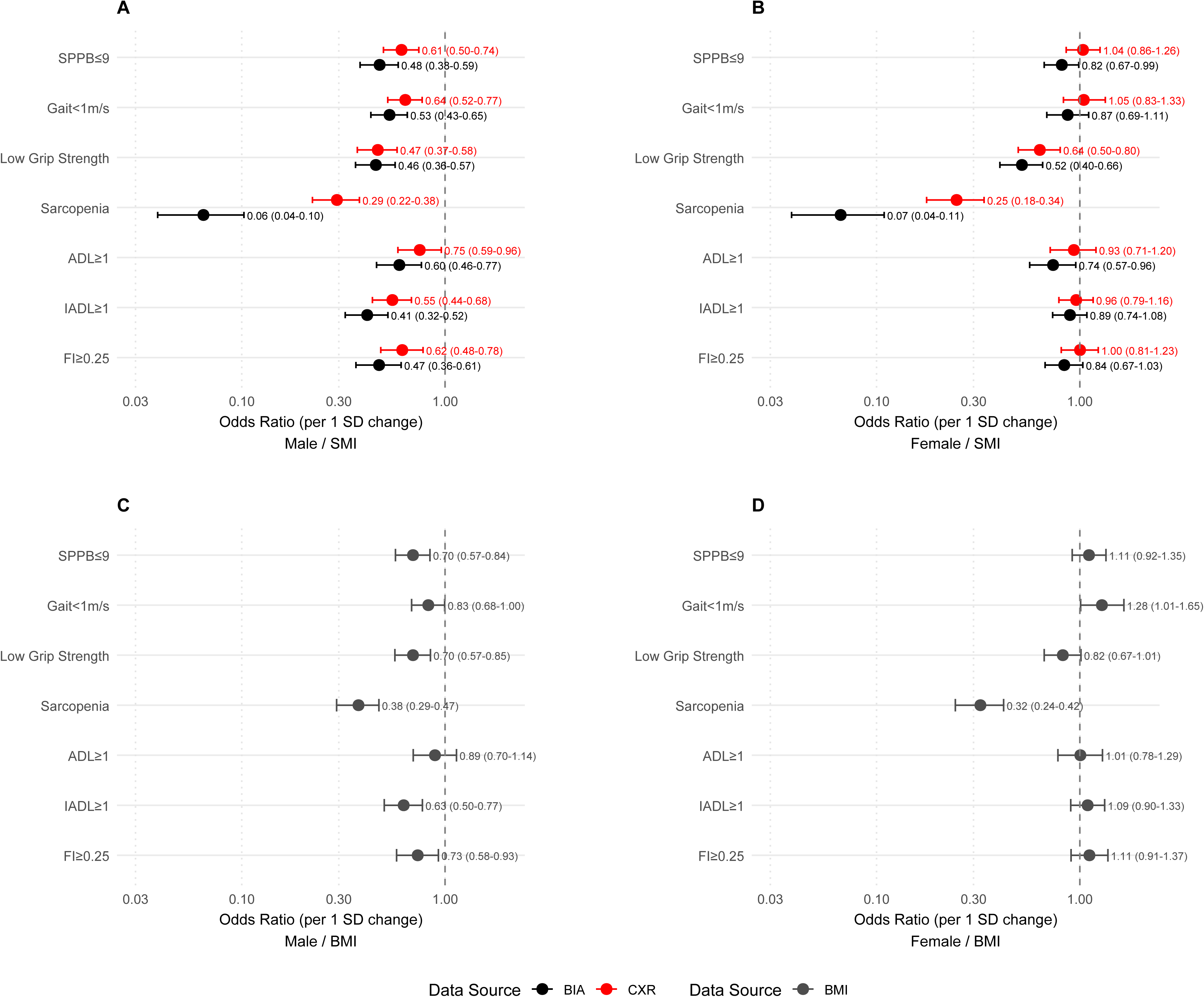
Association between skeletal muscle index and geriatric parameters in the CGA cohort **Note:** Forest plots showing odds ratios (ORs) per 1 SD increase in skeletal muscle index (SMI) for physical performance, gait speed, grip strength, sarcopenia, activities of daily living (ADL) disability, instrumental ADL (IADL) disability, and frailty. Error bars represent 95% confidence intervals (CIs).

## Discussion

A deep-learning model was successfully developed to predict body composition, specifically SMM and FM, from CXRs. The model accurately estimated SMM and FM in the temporal validation, demonstrating strong correlations with BIA–measured values Moreover, the predicted body composition parameters were significantly associated with 10-year all-cause mortality and functional outcomes in older adults, showing patterns comparable to those derived from directly measured data and superior discriminative ability compared with BMI in the independent validation dataset. These findings accentuate not only the predictive accuracy of the model but also its clinical relevance as a potential tool for opportunistic assessment of systemic health using a widely available imaging modality.

To the best of our knowledge, this is the first study to propose a deep-learning model for predicting body composition using CXRs and validate its clinical relevance. Traditionally, CXRs have been used to assess lung fields and cardiac shadows. However, recent studies have demonstrated that CXRs can be used to predict biological age,^34^ as well as various medical conditions such as osteoporosis^21^ and cardiovascular risks.^35^ Theoretically, CXRs incorporate not only the lung and heart but also soft tissues, such as fat and muscles in the chest and parts of the upper extremities and upper abdomen. Our findings indicate that CXRs can be used to predict SMM and FM. It is important to note that the proposed model may capture systemic conditions rather than just muscle or fat mass alone. Body composition reflects systemic conditions such as heart function,^36^ dietary patterns and physical function,^37^ and frailty.^38^

The prognostic utility of the proposed model is notable, and it can be employed in clinical practice as a global health indicator. Body composition is a well-known predictor of health status and mortality.^39^ We demonstrated that the body composition predicted using our model can be used to predict 10-year mortality, and the performance is comparable to actual measurements using BIA. Furthermore, our finding suggests CXR-based body composition, especially SMM, was associated with multiple geriatric parameters, including frailty, physical performance, or disability. Because chest radiography is one of the most basic examinations used for health screening and monitoring disease progression, the proposed model can automatically estimate patients’ body composition from routine CXRs. Furthermore, CXR is commonly utilized for deep-learning algorithms; therefore, the proposed model can be easily employed in clinical practice.^40^ Additionally, as body composition reflects both nutritional and physical health status^41^ and can serve as a prognostic indicator of various diseases,^42^ our model has significant potential for clinical application.

Additionally, its potential applications transcend clinical practice. As it was trained on approximately 100,000 sets of body composition, which is a surrogate marker of general health conditions,^34–37,42^ it can serve as a baseline for developing additional models. For example, it could be used for opportunistic screening of sarcopenia or frailty or predicting the prognosis of other diseases, such as cardiopulmonary or malignant conditions. Furthermore, because CXRs are widely used in both clinical and research settings, our model could help clarify the significance of body composition across different contexts and diseases.

Previous studies have documented the “obesity paradox” in older adults—wherein a BMI is paradoxically linked with lower mortality risk. However, many of these investigations also noted that BMI fails to distinguish between lean muscle and fat mass, thereby limiting its prognostic utility.^43^ In contrast, our study demonstrates that CXR-predicted body composition metrics (lower skeletal muscle index and higher fat percentage) not only replicate the associations observed with direct BIA measurements but also outperform BMI in discriminative ability. By using imaging-derived estimates that specifically separate muscle and fat compartments, our findings suggest a more physiologically meaningful explanation for mortality risk in the older adults.

However, this study has several limitations. First, the proposed model was trained on data from a single health-screening center consisting of relatively healthy individuals, which may limit its generalizability. Although the training and validation datasets were obtained from the same institution, we performed both technical and clinical validation on temporally distinct and independently collected data. Nonetheless, external validation using datasets from other centers is warranted to confirm the robustness and applicability of our findings. Second, the measured BIA values can be susceptible to errors owing to variations in the internal fluid balance.^44^ However, the BIA values employed in this study were reliable as they were measured after the individuals had fasted for at least 8 h. Third, the mortality outcome incidence was low owing to the relatively healthy study population and short follow-up durations. Therefore, we will conduct studies with longer follow-up periods in the future.

## Conclusions

In conclusion, this study demonstrated that CXRs, one of the most widely available medical imaging modalities, can be used to estimate skeletal muscle mass and fat mass through deep learning. The proposed model showed strong agreement with BIA-based measurements and captured clinically meaningful associations with functional status and long-term mortality in both clinically and temporally external datasets. These findings suggest that CXR-based body composition analysis provides an accessible, cost-effective, and noninvasive approach for assessing overall health and frailty in both clinical and population settings.

## Supporting information

CLAIM Checklist

STROPE Checklist

## Author Contribution

Sunghwan Ji and Kiduk Kim contributed equally to this work, including study conceptualization, data interpretation, and manuscript drafting. Kyungjin Cho performed data preprocessing and supported model development. Il-Young Jang and Ji Yeon Baek contributed to the clinical interpretation and provided expertise in geriatric assessment. Miso Jang supervised the technical aspects of model development and validation. Hong-Kyu Kim and Namkug Kim supervised the overall study design, provided critical revisions to the manuscript, and approved the final version. All authors reviewed and approved the submitted manuscript.

## Data Availability Statement

The datasets generated and analyzed during the current study are not publicly available due to institutional and ethical restrictions involving patient privacy. De-identified data may be made available from the corresponding author upon reasonable request and with appropriate approval from the Institutional Review Board.

