## Supplementary material for "Predicting Body Composition from Chest Radiographs by Deep Learning: 10-year Mortality and Geriatric Outcomes": CLAIM Checklist

**Checklist for Artificial Intelligence in Medical Imaging (CLAIM): 2024 Update**

| Section / Topic | No. | Item | Page / Line | No | NA |
| --- | --- | --- | --- | --- | --- |
| TITLE / ABSTRACT |  |  |  |  |  |
|  | **1** | Identification as a study of AI methodology, specifying the category of technology used (e.g., deep learning) | **Title** |  |  |
| ABSTRACT |  |  |  |  |  |
|  | **2** | Summary of study design, methods, results, and conclusions | **Abstract** |  |  |
| INTRODUCTION |  |  |  |  |  |
|  | **3** | Scientific and/or clinical background, including the intended use and role of the AI approach | **Introduction, 3^rd^ paragraph** |  |  |
|  | **4** | Study aims, objectives, and hypotheses | **Introduction, 4^th^ paragraph** |  |  |
| METHODS |  |  |  |  |  |
| *Study Design* | **5** | Prospective or retrospective study | **Methods, 1st paragraph** |  |  |
|  | **6** | Study goal | **Methods, Study Design and Population** |  |  |
| *Data* | **7** | Data sources | **Figure 1** |  |  |
|  | **8** | Inclusion and exclusion criteria | **Figure 1** |  |  |
|  | **9** | Data pre-processing | **Methods, Model Development** |  |  |
|  | **10** | Selection of data subsets | **Figure 1** |  |  |
|  | **11** | De-identification methods | **Methods, Model Development** |  |  |
|  | **12** | How missing data were handled |  |  | **V** |
|  | **13** | Image acquisition protocol | **Methods, Study Design and Population** |  |  |
| *Reference Standard* | **14** | Definition of method(s) used to obtain reference standard | **Methods, Body-composition Measurement** |  |  |
|  | **15** | Rationale for choosing the reference standard | **Methods, Body-composition Measurement** |  |  |
|  | **16** | Source of reference standard annotations | **Methods, Body-composition Measurement** |  |  |
|  | **17** | Annotation of test set |  |  | **V** |
|  | **18** | Measures of inter- and intra-rater variability of features described by the annotators |  |  | **V** |
| *Data Partitions* | **19** | How data were assigned to partitions | **Methods, Study Design and Population** |  |  |
|  | **20** | Level at which partitions are disjoint | **Methods, Study Design and Population** |  |  |
| *Testing Data* | **21** | Intended sample size |  |  | **V** |

| Section / Topic | No. | Item | Page / Line | No | NA |
| --- | --- | --- | --- | --- | --- |
| *Model* | **22** | Detailed description of model | **Methods, Model Development** |  |  |
|  | **23** | Software libraries, frameworks, and packages | **Methods, Model Development&Statistical Analysis** |  |  |
|  | **24** | Initialization of model parameters | **Methods, Model Development** |  |  |
| *Training* | **25** | Details of training approach | **Methods, Model Development** |  |  |
|  | **26** | Method of selecting the final model | **Methods, Model Development** |  |  |
|  | **27** | Ensembling techniques |  |  | **V** |
| *Evaluation* | **28** | Metrics of model performance | **Methods, Statistical Analysis** |  |  |
|  | **29** | Statistical measures of significance and uncertainty | **Methods, Statistical Analysis** |  |  |
|  | **30** | Robustness or sensitivity analysis | **Methods, Statistical Analysis (analysis by subgroup & external validation)** |  |  |
|  | **31** | Methods for explainability or interpretability | **Figure S2** |  |  |
|  | **32** | Evaluation on internal data | **Methods, Statistical Analysis** |  |  |
|  | **33** | Testing on external data | **Methods, Statistical Analysis** |  |  |
|  | **34** | Clinical trial registration |  |  | **V** |
| RESULTS |  |  |  |  |  |
| *Data* | **35** | Numbers of patients or examinations included and excluded | **Results, Baseline Characteristics** |  |  |
|  | **36** | Demographic and clinical characteristics of cases in each partition | **Table 1** |  |  |
| *Model performance* | **37** | Performance metrics and measures of statistical uncertainty | **Table 2** |  |  |
|  | **38** | Estimates of diagnostic performance and their precision |  |  | **V** |
|  | **39** | Failure analysis of incorrect results |  |  | **V** |
| DISCUSSION |  |  |  |  |  |
|  | **40** | Study limitations | **Discussions, 6^th^ paragraph** |  |  |
|  | **41** | Implications for practice, including intended use and/or clinical role | **Discussions, 3rd paragraph** |  |  |
| OTHER INFORMATION |  |  |  |  |  |
|  | **42** | Provide a reference to the full study protocol or to additional technical details |  |  | **V** |
|  | **43** | Statement about the availability of software, trained model, and/or data | **Data Availability Statement** |  |  |
|  | **44** | Sources of funding and other support; role of funders | **Funding** |  |  |

* Indicate page and/or line number for each checklist item that is present. NA = not applicable.
