## Supplementary Materials for "Predicting Body Composition from Chest Radiographs by Deep Learning: 10-year Mortality and Geriatric Outcomes"

**Contents**

- **Method S1**. Model Details

**Figure S1.** Correlation Plots for BIA-measured and Chest Radiograph (CXR)-predicted Skeletal Muscle Mass (SMM) and Fat Mass (FM) for the Validation Dataset

- **Figure S2.** Average Grad-CAM maps for CXR-based prediction of skeletal muscle mass (SMM) and fat percentage by sex.
- **Figure S3.** Restricted cubic spline plots showing hazard ratios (HRs) of 10-year all-cause mortality according to skeletal muscle index, fat percentage, and body mass index
- **Figure S4.** Odds ratios per 1 standard deviation increase in fat percentage for geriatric parameters, comparing BIA- and CXR-derived estimates in men (A) and women (B)
- **Table S1.** 50-item frailty index
- **Table S2.** Mean Absolute Errors (MAEs) and Correlation Coefficients (*r*) For Predicted Fat Mass (FM), Skeletal Muscle Mass (SMM), Age, Height, Weight, and Sex for the Temporal Validation Dataset (≥65 years)
- **Table S3.** Mean Absolute Errors (MAE) and Correlation Coefficients (*r*) of the Skeletal Muscle Mass (SMM) Model for Predicting Fat Mass, Age, Height, Weight, and Sex According to Sex.
- **Table S4.** Mean Absolute Errors (MAE) and Correlation Coefficients (r) of the Fat Mass (FM) Model for Predicting Fat Mass, Age, Height, Weight, and Sex According to Sex.
- **Table S5.** Concordance measures between BIA-based and CXR-based HR estimates for all-cause mortality
- **Table S6.** Hazard Ratios (HRs) of Mortality According to Tertiles of Measured and Predicted Fat Percentages

**Method S1.** Model Details

In this study, a label-distribution learning strategy with additional mean-variance loss^34^ was used to predict continuous variables from CXRs. Therefore, regression tasks for predicting continuous variables were transferred to classification tasks. The cross-entropy loss between the gold-standard class label and the predicted output can then be calculated as follows:

$L_{CE}=-\frac{1}{N}\sum_{t=1}^{N} i_{t}*\log p_{x,t}$	(1)

where $i_{t}\in\{i_{1},i_{2},\ldots,i_{t},\ldots,i_{N}\}$ denotes the class labels (i.e., possible values of continuous variables) and $p_{x}$ denotes the estimated class distribution for sample *x* over *N* classes. Thus, $p_{x,t}$ denotes the probability that sample *x* belongs to class $i_{t}$. Additionally, the softmax output of a deep-learning model can be considered a probability density function for the label distribution. Therefore, the expectation of the predicted output $E\left( x \right)$ and variance $Var\left( x \right)$ can be calculated as follows:

$E\left( x \right)=\sum_{t=1}^{N} i_{t}*p_{x,t}$	(2)

$Var\left( x \right)=\sum_{t=1}^{N} p_{x,t}*{(i_{t}-E(x))}^{2}$	(3)

Finally, the mean loss was calculated to minimize the difference between the gold-standard labels and the expectation of the predicted output, and the variance was minimized as a loss function as follows:

$L_{mean}={(E\left( x \right)-i_{x})}^{2}={(\sum_{t=1}^{N} (i_{t}*p_{x,t})-i_{x})}^{2}$	(4)

$L_{var}=Var\left( x \right)=\sum_{t=1}^{N} p_{x,t}*{(i_{t}-\sum_{j=1}^{N} i_{j}*p_{x,j})}^{2}$	(5)

Accordingly, the final loss was calculated as the sum of the individual losses as follows:

$Loss=L_{CE}+L_{mean}+L_{var}$	(6)

All models were trained from scratch with random initialization. For training, one graphic processing unit of Titan RTX from NVIDIA with a batch size of 64 was used. AdamW without scheduler was used for optimization. The learning rate was set at 0.0003 (3e-4). All models were implemented with the PyTorch version of 2.1.0.

**Figure S1.** Correlation Plots for BIA-measured and Chest Radiograph (CXR)-predicted Skeletal Muscle Mass (SMM) and Fat Mass (FM) for the Validation Dataset: (A) FM for Females; (B) FM for Males; (C) SMM for Females; (D) SMM for Males. BIA, Bioelectrical Impedance Analysis

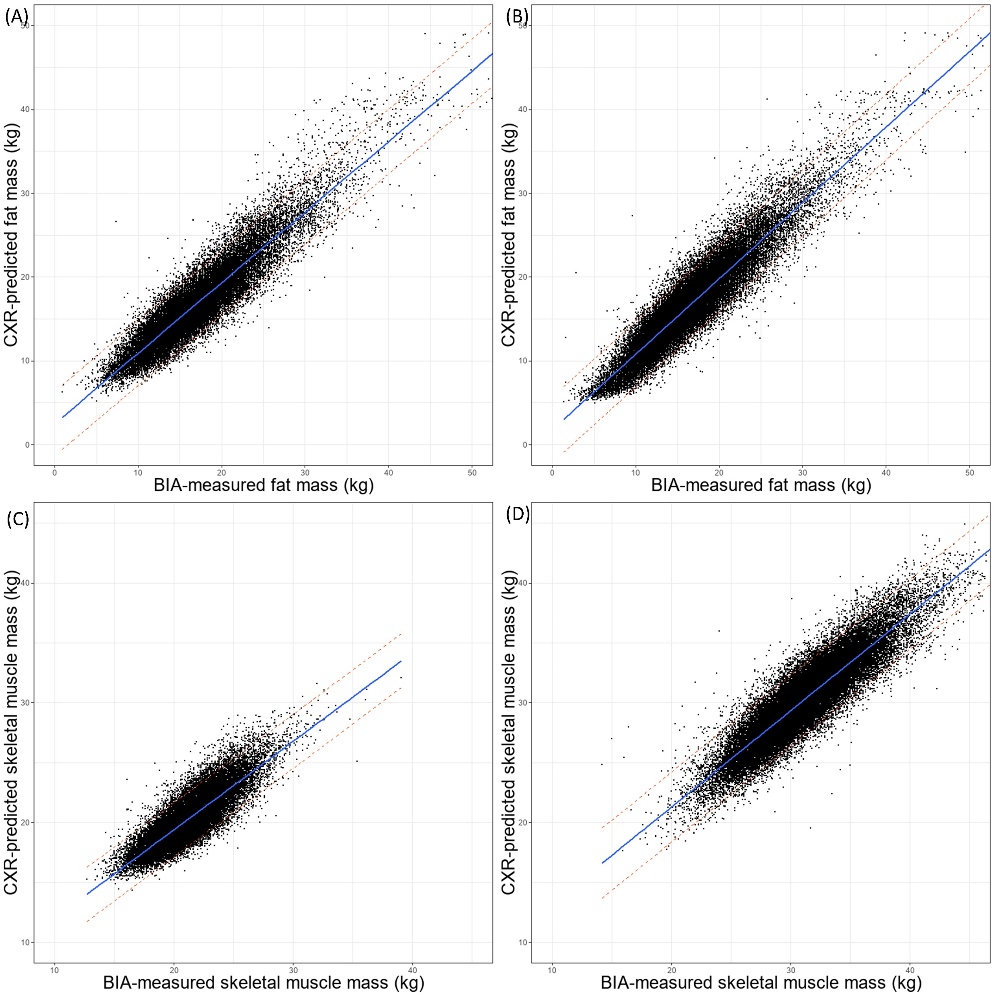

**Figure S2.** Average Grad-CAM maps for CXR-based prediction of skeletal muscle mass (SMM) and fat percentage by sex. (A)Female SMM; (B)Male SMM; (C) Female fat percentage; (D) Male fat percentage.
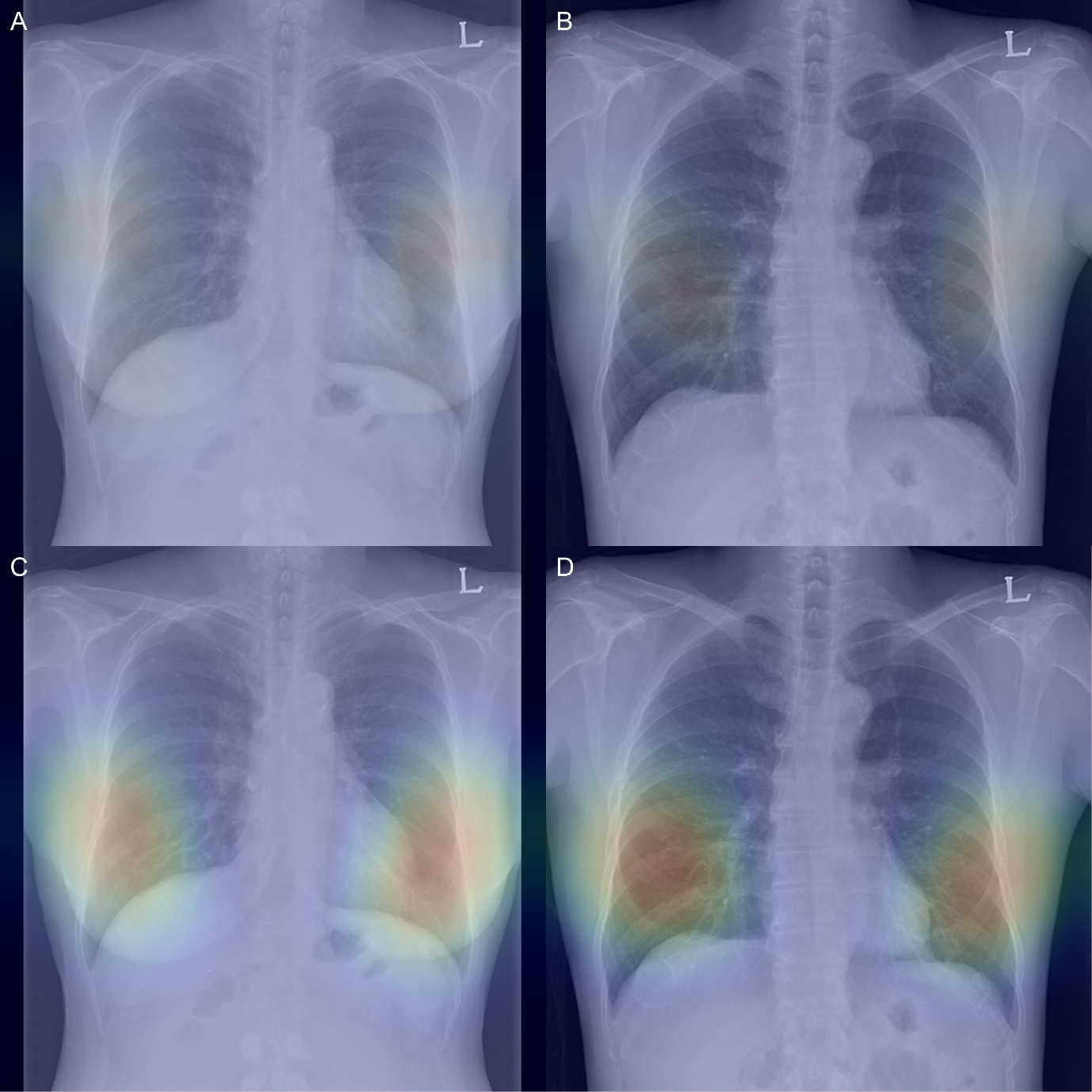

**Figure S3.** Restricted cubic spline plots showing hazard ratios (HRs) of 10-year all-cause mortality according to skeletal muscle index (A, female; B, male), fat percentage (C, female; D, male), and body mass index (E, female; F, male); gray lines represent BIA-measured values and red lines represent CXR-predicted values, with shaded areas indicating 95% confidence intervals.

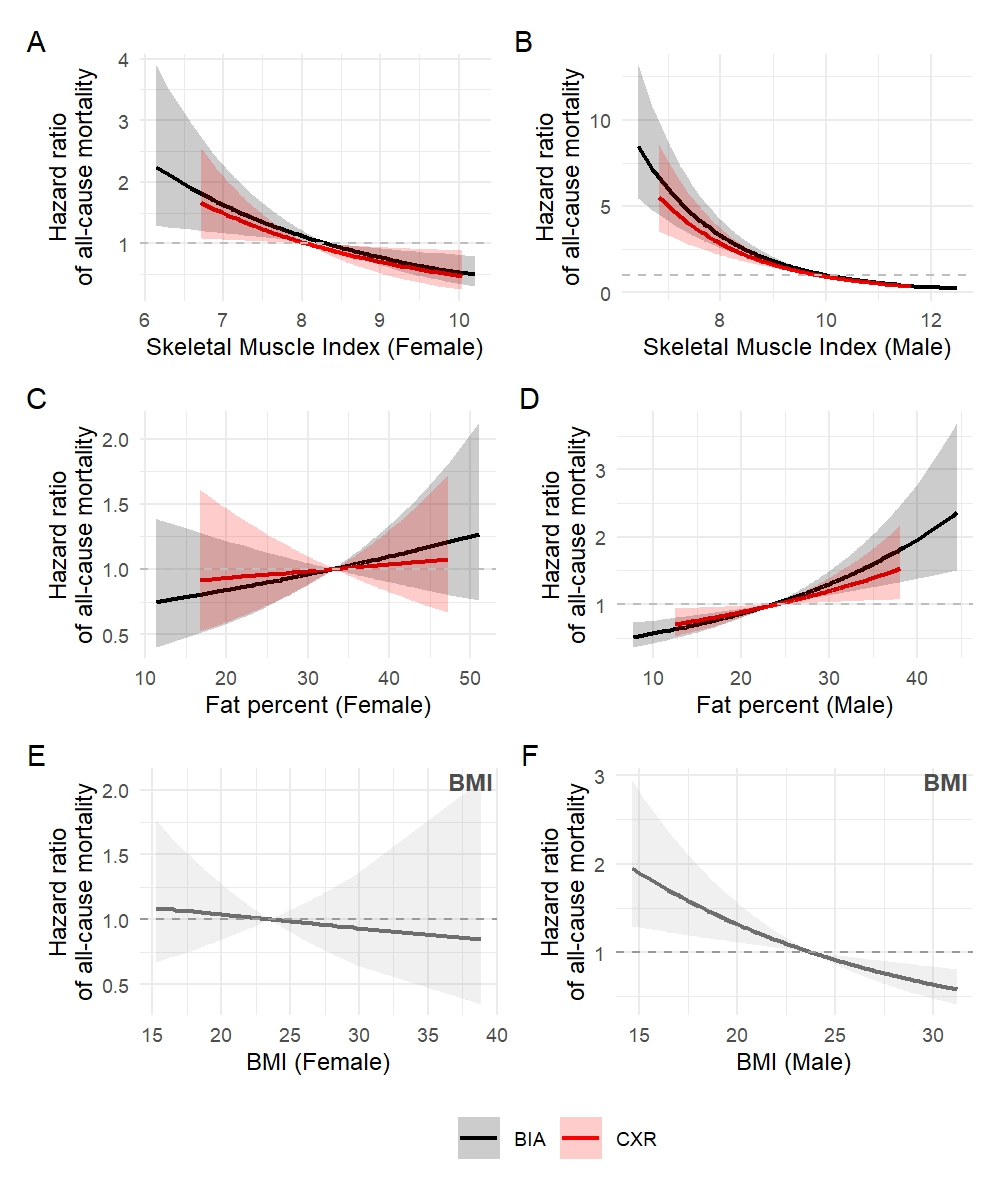

**Figure S4.** Odds ratios per 1 standard deviation increase in fat percentage for geriatric parameters, comparing BIA- and CXR-derived estimates in men (A) and women (B)

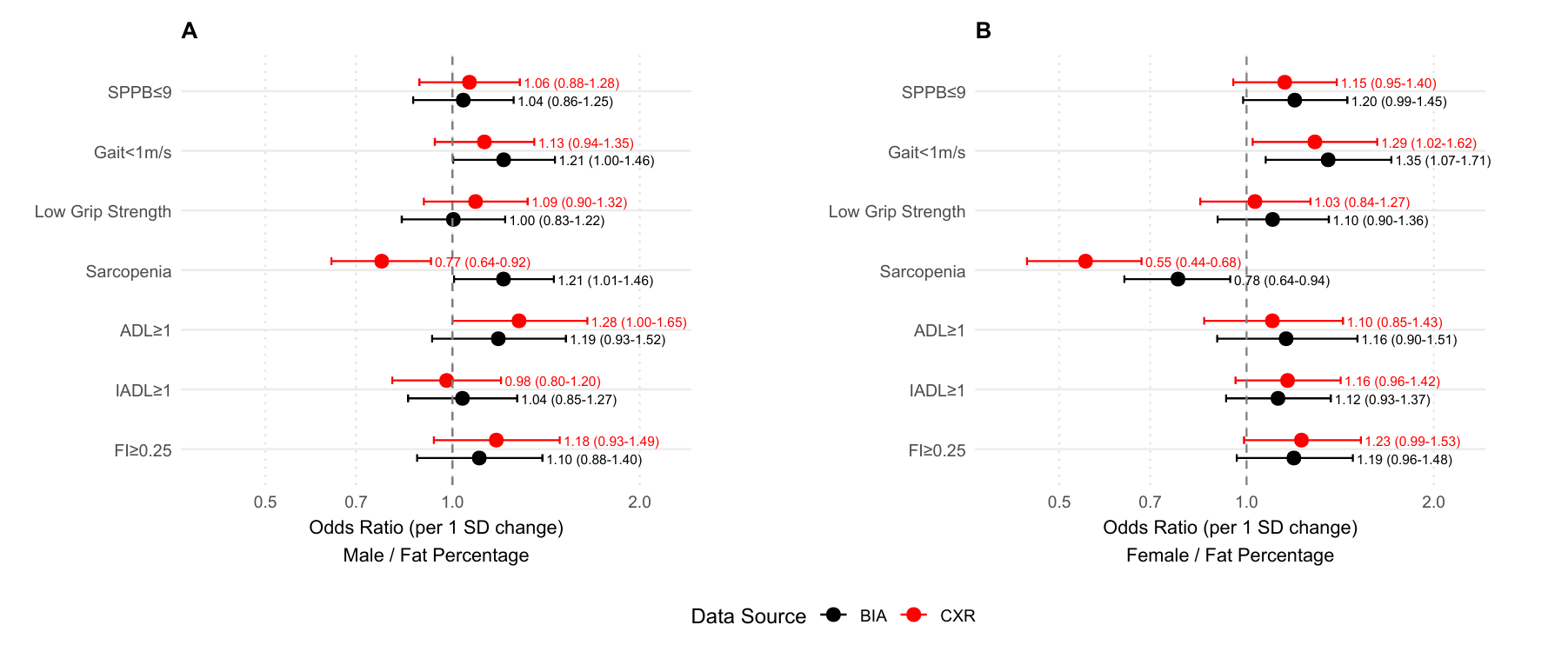

**Table S1.** 50-item frailty index

**Composition of 50-item comprehensive geriatric assessment-frailty index (CGA-FI) used in this study**

**1. Medical history (21 deficit items)**

Hypertension (yes, 1; no, 0)

Diabetes mellitus (yes, 1; no, 0)

Chronic kidney disease (yes, 1; no, 0)

Atrial fibrillation (yes, 1; no, 0)

Angina pectoris (yes, 1; no, 0)

History of myocardial infarction (yes, 1; no, 0)

Congestive heart failure (yes, 1; no, 0)

Peripheral artery disease (yes, 1; no, 0)

Chronic obstructive pulmonary disease (yes, 1; no, 0)

Cerebral artery disease (yes, 1; no, 0)

History of stroke (yes, 1; no, 0)

Dementia (yes, 1; no, 0)

Sensory impairment (yes, 1; no, 0)

Depression (yes, 1; no, 0)

Anxiety disorder (yes, 1; no, 0)

Malignant disease (yes, 1; no, 0)

Arthritis (yes, 1; no, 0)

Spinal disease (yes, 1; no, 0)

Asthma (yes, 1; no, 0)

Fall history within 1 year (yes, 1; no, 0)

Polypharmacy (≥5 medications; yes, 1; no, 0)

**2. Functional status (22 deficit items)**

***Activities of Daily Living**

Difficulties in dressing (yes, 1; no, 0)

Difficulties in grooming (yes, 1; no, 0)

Difficulties in bathing (yes, 1; no, 0)

Difficulties in eating (yes, 1; no, 0)

Difficulties in moving indoors (yes, 1; no, 0)

Difficulties in getting in and out of bed (yes, 1; no, 0)

Difficulties in toileting (yes, 1; no, 0)

***Instrumental Activities of Daily Living**

Difficulties in using phone (yes, 1; no, 0)

Difficulties in buying groceries (yes, 1; no, 0)

Help needed in transportation (yes, 1; no, 0)

Help needed in managing medications (yes, 1; no, 0)

Difficulties in managing finances (yes, 1; no, 0)

Difficulties in preparing foods (yes, 1; no, 0)

Difficulties in performing basic household chores (yes, 1; no, 0)

***Nagi & Rosow-Breslau activities**

Difficult to perform stooping, crouching, or kneeling (yes, 1; no, 0)

Difficult to lift or carry objects as heavy as 10 pounds (yes, 1; no, 0)

Difficult to write or handle and grasp small objects (yes, 1; no, 0)

Difficult to pull or push large objects (yes, 1; no, 0)

Difficult to reach or extend arms above the shoulder level (yes, 1; no, 0)

Help needed to do heavy work around the house (yes, 1; no, 0)

Help needed to walk upstairs and downstairs (yes, 1; no, 0)

Help needed to walk half a mile (yes, 1; no, 0)

**3. Performance tests (four deficit items)**

Hand grip strength, kg (0 for ≥32, 0.5 for ≥26 and <32, 1 for <26 in men; 0 for ≥20, 0.5 for ≥16 and <20, 1 for <16 in women)

Usual gait speed, m/s (0 for ≥1, 0.3 for ≥0.8 and <1, 0.7 for ≥0.6 and <0.8, 1 for <0.6)

Chair rise test time, s (0 for <11.2, 0.25 for ≥11.2 and <13.7, 0.5 for ≥13.7 and <16.7, 0.75 for ≥16.7 and <61, 1 for ≥61)

MMSE score (0.3 for 24–26, 0.7 for 21–23, 1 for 0–20)

**4. Nutritional status (three deficit items)**

Weight loss >4.5 kg within 1 year (yes, 1; no, 0)

Body mass index <18.5 kg/m^2^ (yes, 1; no, 0)

Serum albumin level <3.5 g/dL (yes, 1; no, 0)

MMSE, Mini–Mental Status Examination

**Table S2.** Mean Absolute Errors (MAEs) and Correlation Coefficients (*r*) For Predicted Fat Mass (FM), Skeletal Muscle Mass (SMM), Age, Height, Weight, and Sex for the Temporal Validation Dataset (≥65 years)

|  | | **Temporal validation dataset (≥65 years)  (Females N = 2,431, Males N = 3,501)** | |
| --- | --- | --- | --- |
|  |  | **MAE (mean±SD)** | **r (95% confidence interval)** |
| SMM (kg) | | 1.43 ± 1.13 | 0.944 (0.941–0.946) |
| FM (kg) | | 1.75 ± 1.45 | 0.910 (0.905–0.914) |
| Age (years) | FM model | 4.18 ± 3.00 | 0.662 (0.647–0.676) |
|  | SMM model | 3.76 ± 2.78 | 0.659 (0.644–0.673) |
| Height (m) | FM model | 0.02 ± 0.02 | 0.929 (0.925–0.932) |
|  | SMM model | 0.03 ± 0.02 | 0.926 (0.922–0.929 |
| Weight (kg) | FM model | 2.50 ± 1.99 | 0.953 (0.951–0.955) |
|  | SMM model | 2.58 ± 2.03 | 0.952 (0.949–0.954) |
| Sex (Accuracy %) | FM model | 99.7 | |
|  | SMM model | 99.7 | |

Note: N corresponds to the number of chest radiographs. MAE, mean absolute error; SD, standard deviation.

**Table S3.** Mean Absolute Errors (MAE) and Correlation Coefficients (*r*) of the Skeletal Muscle Mass (SMM) Model for Predicting Fat Mass, Age, Height, Weight, and Sex According to Sex.

|  | **Validation dataset (N = 77,655)** | | | | **Temporal validation dataset (≥65 years) (N = 5,932)** | | | |
| --- | --- | --- | --- | --- | --- | --- | --- | --- |
|  | **Females (N = 33,085)** | | **Males (N = 44,570)** | | **Females (N = 2,431)** | | **Males (N = 3,501)** | |
|  | **MAE** | ***r*** | **MAE** | ***r*** | **MAE** | ***r*** | **MAE** | ***r*** |
| Skeletal Muscle Mass (kg) | 1.29 ± 0.97 | 0.844 (0.840–0.847) | 1.48 ± 1.19 | 0.899 (0.897–0.901) | 1.25 ± 0.96 | 0.791 (0.776–0.805) | 1.55 ± 1.22 | 0.833 (0.823–0.843) |
| Age (years) | 2.76 ± 2.19 | 0.942 (0.941–0.944) | 2.85 ± 2.29 | 0.933 (0.932–0.934) | 3.56 ± 2.59 | 0.671 (0.648–0.692) | 3.89 ± 2.91 | 0.653 (0.634–0.672) |
| Height (m) | 0.02 ± 0.02 | 0.872 (0.870–0.875) | 0.02 ± 0.02 | 0.864 (0.862–0.867) | 0.02 ± 0.02 | 0.811 (0.797–0.824) | 0.03 ± 0.02 | 0.794 (0.782–0.806) |
| Weight (kg) | 2.50 ± 1.97 | 0.938 (0.937–0.939) | 2.40 ± 2.04 | 0.959 (0.958–0.960) | 2.69 ± 2.12 | 0.917 (0.910–0.923) | 2.51 ± 1.96 | 0.939 (0.935–0.943) |
| Sex (accuracy, %) | 99.94 | | 99.85 | | 99.63 | | 99.74 | |

N: Number of Chest Radiographs.

**Table S4.** Mean Absolute Errors (MAE) and Correlation Coefficients (*r*) of the Fat Mass (FM) Model for Predicting Fat Mass, Age, Height, Weight, and Sex According to Sex.

|  | **Validation dataset (N = 77,655)** | | | | **Temporal validation dataset (≥65 years) (N = 5,932)** | | | |
| --- | --- | --- | --- | --- | --- | --- | --- | --- |
|  | **Females (N = 33,085)** | | **Males (N = 44,570)** | | **Females (N = 2,431)** | | **Males (N = 3,501)** | |
|  | **MAE** | ***r*** | **MAE** | ***r*** | **MAE** | ***r*** | **MAE** | ***r*** |
| Fat Mass (kg) | 1.67 ± 1.38 | 0.923 (0.921–0.924) | 1.57 ± 1.36 | 0.926 (0.924–0.927) | 1.87 ± 1.57 | 0.903 (0.896–0.910) | 1.66 ± 1.35 | 0.905 (0.898–0.910) |
| Age (years) | 2.84 ± 2.27 | 0.939 (0.938–0.941) | 2.78 ± 2.26 | 0.932 (0.930–0.933) | 4.23 ± 2.93 | 0.666 (0.643–0.687) | 4.15 ± 3.04 | 0.660 (0.641–0.678) |
| Height (cm) | 2 ± 2 | 0.872 (0.869–0.874) | 2 ± 2 | 0.867 (0.864–0.869) | 2 ± 2 | 0.819 (0.805–0.832) | 3 ± 2 | 0.801 (0.788–0.812) |
| Weight (kg) | 2.31 ± 1.89 | 0.941 (0.940–0.942) | 2.40 ± 2.03 | 0.957 (0.956–0.958) | 2.48 ± 2.03 | 0.924 (0.918–0.930) | 2.51 ± 1.96 | 0.939 (0.935–0.943) |
| Sex (accuracy, %) | 99.95 | | 99.80 | | 99.96 | | 99.46 | |

N, Number of Chest Radiographs.

**Table S5.** Concordance measures between BIA-based and CXR-based HR estimates for all-cause mortality

| Panel | Pearson *r* | CCC |
| --- | --- | --- |
| SMI (Female) | 1.00 | 0.98 |
| SMI (Male) | 1.00 | 0.97 |
| Fat percent (Female) | 1.00 | 0.68 |
| Fat percent (Male) | 1.00 | 0.91 |
